# Vitamin D status, body mass index, ethnicity and COVID-19: Initial analysis of the first-reported UK Biobank COVID-19 positive cases (*n* 580) compared with negative controls (*n* 723)

**DOI:** 10.1101/2020.04.29.20084277

**Authors:** Andrea L. Darling, Kourosh R. Ahmadi, Kate A Ward, Nicholas C Harvey, Alexessander Couto Alves, Deborah K. Dunn-Walters, Susan A. Lanham-New, Cyrus Cooper, David J. Blackbourn

## Abstract

*In this short report we present a preliminary assessment of the serum 25-hydroxyvitamin D status (25(OH)D), body mass index (BMI), ethnicity and other lifestyle factors in the first-reported UK Biobank COVID-19 positive cases (n 580) compared with negative controls (n 723)*. The COVID-19 cases include those who have been treated as a hospital inpatient as well as those who have not, and are from England only. Mean (SD) for age was 57.5 (8.7) in positive cases and 57.9 (8.7) in negative controls.

---

*Serum 25(OH)D status was almost identical in those who tested positive (median (Interquartile range, IQR)= 43.3 (32.1) nmol/L) compared to those who tested negative (median (IQR) 44.1 (31.2) nmol/L) for COVID-19*. It must be borne in mind that the sample on average was not severely vitamin D deficient and results may differ in populations with a higher prevalence of severe vitamin D deficiency. As shown in Table 1, there was no difference in 25(OH)D status by gender; however 25(OH)D was significantly lower in those with obesity [P<0.001] by 9 nmol/L compared with those of normal or overweight. Of note, 25(OH)D status was also significantly lower in those of Asian, Black and Mixed ethnicity (by 16nmol/L) [P<0.001] compared with those of White ethnicity. This supports previous findings of a low serum 25(OH)D in UK South Asian individuals from the UK DFINES study^1^.

**Table 1:** Serum 25(OH)D status by participant characteristic and COVID-19 test result

| 25(OH)D Median (IQR) nmol/L | Gender |  | Body Mass Index (BMI) |  |  | Ethnicity |  | Mann-Whitney/Kruskal Wallis test |  |  |
| --- | --- | --- | --- | --- | --- | --- | --- | --- | --- | --- |
|  | Female | Male | Normal/under weight | Overweight | Obesity | Asian, Black, Mixed and Other | White | P for gender | P for BMI | P for ethnic |
| All | 42.9 (31.7), n=590 | 44.1 (31.1), n=713 | 48.0 (32.4), n=378 | 47.0 (31.5), n=441 | 38.5 (28.4), n=468 | 30.0 (22.5), n=134 | 46.0 (31.5), n=1160 | 0.34 | <0.001 | <0.001 |
| Cases (+ve) | 41.7 (31.1), n=244 | 44.9 (32.3), n=336 | 47.1 (32.6), n=135 | 46.3 (31.3), n=208 | 38.6 (29.4), n=231 | 28.5 (22.2), n=81 | 46.0 (31.0), n=494 | 0.24 | 0.001 | <0.001 |
| Controls (-ve) | 44.2 (32.4), n=346 | 43.6 (30.7), n=377 | 48.5 (31.7), n=243 | 47.5 (31.1), n=233 | 38.2 (27.1), n=237 | 32.3 (21.8), n=53 | 46.0 (31.4), n=666 | 0.81 | <0.001 | <0.001 |

*A logistic regression model (Table 2) suggests that being overweight or obese; living in London; being male and being of Asian, Black or Mixed ethnicity is associated with a higher odds of testing positive for COVID-19, which confirms clinical observations which have been reported in the media*.

**Table 2:** Odds ratio of a positive COVID-19 test result-n=561 positive cases and n=705 negative controls-preliminary logistic regression model

| Model P<0.001<br>Nagelkerke R<br>Square=0.06 | Participant<br>n | Beta | Standard<br>Error | P value | Odds ratio<br>(OR) | 95% C.I.for<br>OR lower | 95% C.I.for<br>OR upper |
| --- | --- | --- | --- | --- | --- | --- | --- |
| Townsend Deprivation<br>Index below median<br>(lower deprivation) | 646 |  |  |  | 1.00 |  |  |
| Townsend Deprivation<br>Index above median<br>(higher deprivation) | 620 | 0.05 | 0.12 | 0.70 | 1.05 | 0.82 | 1.34 |
| **Region: England-**<br>**Outside London** | 1031 |  |  |  | **1.00** |  |  |
| **Region: England-**<br>**London** | 235 | **0.37** | **0.16** | **0.02** | **1.45** | **1.05** | **2.00** |
| Self-reported health:<br>Excellent/good | 734 |  |  |  | 1.00 |  |  |
| Self-reported health:<br>Fair/poor | 532 | -0.13 | 0.12 | 0.29 | 0.88 | 0.69 | 1.12 |
| **BMI: Normal/Under**<br>**weight** | 375 |  |  |  | **1.00** |  |  |
| **BMI: Overweight** | 433 | **0.41** | **0.15** | **0.01** | **1.51** | **1.13** | **2.02** |
| **BMI: Obesity** | 458 | **0.52** | **0.15** | **0.00** | **1.67** | **1.24** | **2.26** |
| Age category (Baseline<br>age 40-60 y; currently<br>50-70y) | 606 |  |  |  | 1.00 |  |  |
| Age category (Baseline<br>age over 60y; currently<br>over 70 y) | 660 | -0.25 | 0.12 | 0.04 | 0.78 | 0.62 | 0.99 |
| Q1 25(OH)D Bottom<br>25% | 314 |  |  |  | 1.00 |  |  |
| Q2 25(OH)D | 317 | -0.08 | 0.17 | 0.65 | 0.93 | 0.67 | 1.28 |
| Q3 25(OH)D | 314 | 0.03 | 0.17 | 0.84 | 1.03 | 0.74 | 1.44 |
| Q4 (24(OH)D Top 25% | 321 | 0.11 | 0.17 | 0.54 | 1.11 | 0.79 | 1.56 |
| **Ethnicity: White** | 1139 |  |  |  | **1.00** |  |  |
| **Ethnicity: Asian, Black,**<br>**Mixed and Other** | 127 | **0.51** | **0.22** | **0.02** | **1.66** | **1.08** | **2.54** |
| **Smoking: Non smoker** | 1080 |  |  |  | **1.00** |  |  |
| **Smoking: Regular**<br>**Smoker** | 142 | **-0.54** | **0.20** | **0.01** | **0.58** | **0.39** | **0.86** |
| Smoking: Occasional | 44 | -0.19 | 0.32 | 0.56 | 0.83 | 0.44 | 1.56 |
| **Gender: Female** | 579 |  |  |  | **1.00** |  |  |
| **Gender: Male** | 687 | **0.24** | **0.12** | **0.04** | **1.28** | **1.01** | **1.61** |
| Constant |  | -0.61 | 0.19 | 0.00 | 0.55 |  |  |

Variables in bold face are statistically significant based on 95% confidence intervals for OR. 25(OH)D= 25-hydroxyvitamin D (nmol/L).
Q1=Quartile1, Q2= Quartile 2, Q3=Quartile 3, Q4=Quartile 4. y=years.

Being a regular smoker (smoking on all or most days) was associated with a reduced odds (OR=0.58 (0.39-0.86)) of testing positive, compared with being a non-smoker (OR=1). However a key limitation of this analysis is that there were only a small number of regular smokers (n 142) and persons of Asian, Black or Mixed/Other Ethnicity in the sample and larger populations will be required to confirm these results.

Another limitation of our analysis is that vitamin D status was assessed in the UK Biobank cohort using baseline samples which were collected in 2006–2010. However, there is scientific evidence to show that that an individual’s vitamin D status tends to track over time; for example, those with status in the top quartile of 25(OH)D status in the population will likely still be in that range over a decade later^2^. Therefore, we used quartiles of 25(OH)D status in our analysis rather than the specific 25(OH)D value which showed a null association with Covid-19 test result. When a BMI x 25(OH)D status (continuous variable) interaction term was trialed in the model, it was not statistically significant suggesting the interaction between BMI and 25(OH)D status did not predict test result in the currently available dataset.

Nevertheless, from our initial statistical model of the UK Biobank data, we can conclude a higher odds of testing positive for COVID-19 is associated with the cofactors of living in London; being male and being of Asian, Black or Mixed ethnicity.

As the number of reported cases increases in the UK Biobank, we will expand our model to control for additional factors such as blood pressure, use of statin medications, diagnoses of cardiovascular disease, respiratory disease, diabetes, COVID-19-attributed mortality and conditions affecting immune function, as well as genetics.

This research was conducted under UK Biobank Project 15168; the views are those of the authors’ own. *Note: DKDW is not a collaborator on a UK Biobank project and did not view the data*.

## Data Availability

Data is available from the UK Biobank directly- the authors are not authorised to provide this to others.

## Ethics

The UK Biobank study was conducted according to the guidelines laid down in the Declaration of Helsinki and all procedures involving human subjects were approved by the UK North West Multi-centre Research Ethics Committee (MREC); application 11/NW/0382. Written informed consent was obtained from all subjects.

## References

1 Darling AL, Hart KH, Macdonald HM, Horton K, Kang’ombe AR, Berry JL, Lanham-New SA. Vitamin D deficiency in UK South Asian Women of childbearing age: a comparative longitudinal investigation with UK Caucasian women. Osteoporos Int. 2013: 24:477–88.

2 Jorde R, Sneve M, Hutchinson M, Emaus N, Figenschau Y, Grimnes G. Tracking of serum 25-hydroxyvitamin D levels during 14 years in a population-based study and during 12 months in an intervention study. Am J Epidemiol. 2010: 171:903–8.

